# Estimating gender disparities in surgical sterilization uptake in India in 2019-20 and cost savings from equity achievement

**DOI:** 10.64898/2026.06.05.26354923

**Authors:** Sharvari Mande, Anoushka Arora, Parth Sharma, Varun Raj Passi, Aiman Perween Afsar, Keerty Nakray, Himani Baxy, Siddhesh Zadey

**Affiliations:** Association of Socially Applicable Research (ASAR), Pune, Maharashtra, India; Seth G.S Medical College, Parel, Mumbai; Organization for Research and Education on Social Policy; Independent OBGYN practitioner and researcher; Department of Epidemiology, Mailman School of Public Health, Columbia University, NYC, NY, USA; Dr. D.Y. Patil Dental College and Hospital, Dr. D. Y. Patil Vidyapeeth, Pune, Maharashtra, India; GEMINI Research Center, Duke University School of Medicine, Durham, NC, USA

## Abstract

**Background:** Qualitative studies have noted that the burden of family planning disproportionately falls on women in India. Our primary objective was to quantify the gender disparity in the uptake of surgical sterilizations. Our secondary objectives were to calculate the costs of tubectomies and vasectomies in India and to estimate the savings of scaling up vasectomy rates.

**Methods:** We conducted a retrospective analysis using data on the total number of tubectomies and vasectomies performed, postoperative failure, and postoperative mortality due to these procedures, obtained from the Health Management Information System (HMIS) for 2019-20. We calculated the vasectomy (tubectomy) operative rates per 10,000 men (women) of reproductive age (15-49 years). The women-to-men ratio of these rates is used as a proxy for sex-based disparities in uptake. State-specific procedure costs and compensation for failures and postoperative deaths at public hospitals were extracted and aggregated from government data and research studies. To estimate the financial benefit of scaling up vasectomies, the cost of increasing the vasectomy rate to 50% of the total sterilization rate was calculated. All costs were adjusted for inflation to 2022 and presented in United States Dollars (USD).

**Findings:** In 2019-20, the national tubectomy rate was 96.5, the vasectomy rate was 1.4, and the resulting women-to-men rate ratio was 67.5. The cost per tubectomy procedure was 3.5 times that of vasectomy (89.1 USD vs. 25.3 USD). Keeping the overall operative rate constant, the net savings from scaling up vasectomies to at least 50% of total operations (replacing excess tubectomies) range from 62,193,487 to 75,355,777 USD.

**Interpretation:** Our pan-India analysis confirms that the use of surgical family planning methods is disproportionately higher among women. Scaling up vasectomies has finacial benefits and can improve gender equity.

**Funding:** None.

## 1 Introduction

Gender disparity, limited opportunities for women compared to men, and neglect towards women’s health and wellbeing, except for maternal health, are widespread in India. Such disparities, founded in patriarchal socio-cultural tradition, manifest all the way from the disproportionate burden of unpaid work on women to their limited role in leadership, including health policy leadership, (1) with women’s interests being neglected across the board. (2)

The burden of family planning in India is also gendered. While the unmet need for family planning has decreased from 20.6% in 1993 to 9.4% in 2021, the responsibility has fallen mainly on women. (3) This is highlighted by the fact that, despite being a more invasive procedure as compared to male sterilization, female sterilization (37.9%) continues to be the most commonly used method of contraception in India. (4) Compared to tubectomy (female surgical sterilization), vasectomy (male surgical sterilization) is a more straightforward, less invasive procedure with lower complication rates and failure rates, making it a better option for permanent contraception, particularly for women in long-term single-partner relationships. (5) However, as per the National Family Health Survey (NFHS) 2019-21, only 0.3% of the eligible men underwent the procedure compared to 38% of eligible women. (6)

The gender disparity in the uptake of surgical sterilization and the added costs to the public health system of this disparity remain unknown for India, especially at the level of states and districts where health policymaking and programming are conducted. We aimed to fill this gap. We estimated the gender disparities in the surgical sterilization uptake for Indian districts and states. Furthermore, we estimated the potential savings for the health system if vasectomies are scaled up in India to achieve gender-equitable surgical sterilization rates.

## 2 Methods

### 2.1 Data Sources and Variables

We conducted a retrospective cross-sectional study across 707 districts in 36 states and union territories (UTs) of India for the financial year 2019-20, using publicly available data **(Table 1)**.

**Table 1:**
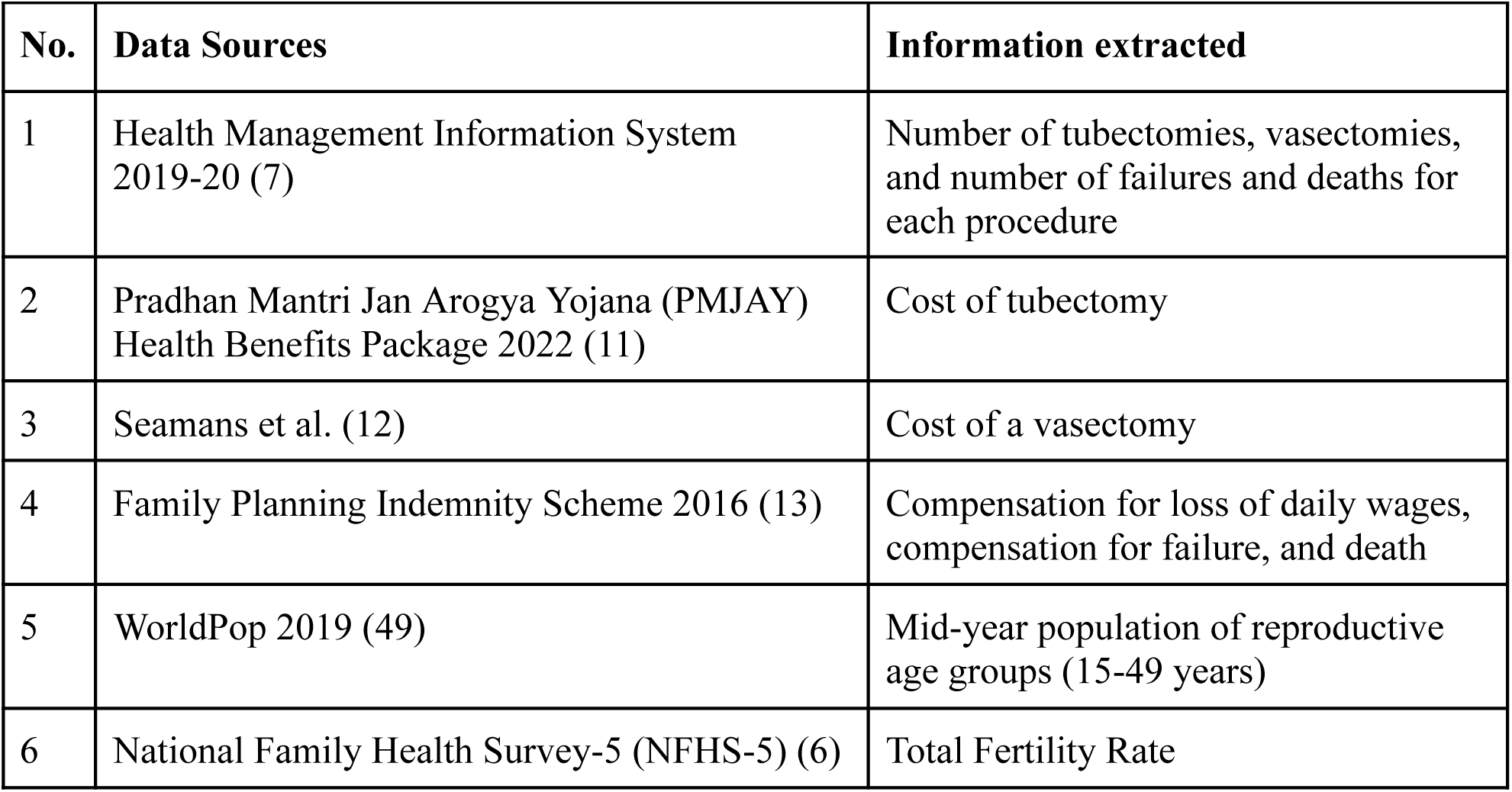
Data sources and variables.

| **No.** | **Data Sources** | **Information extracted** |
| --- | --- | --- |
| 1 | Health Management Information System 2019-20 (7) | Number of tubectomies, vasectomies, and number of failures and deaths for each procedure |
| 2 | Pradhan Mantri Jan Arogya Yojana (PMJAY) Health Benefits Package 2022 (11) | Cost of tubectomy |
| 3 | Seamans et al. (12) | Cost of a vasectomy |
| 4 | Family Planning Indemnity Scheme 2016 (13) | Compensation for loss of daily wages, compensation for failure, and death |
| 5 | WorldPop 2019 (49) | Mid-year population of reproductive age groups (15-49 years) |
| 6 | National Family Health Survey-5 (NFHS-5) (6) | Total Fertility Rate |

We used the Health Management Information System (HMIS) to obtain the total number of tubectomies and vasectomies performed, as well as the number of failures and deaths associated with each procedure. (7) HMIS also provided the number of sterilization procedures that took place in public and private health centers. Due to data availability and policy relevance, we included only tubectomies performed at public health centers in this analysis (see limitations below). We considered two surgical methods of tubectomies: open and laparoscopic. Open tubectomies included procedures conducted post-abortion, post-partum, as well as those utilizing mini-laparotomy techniques. Hence, the total number of tubectomies performed comprised procedures conducted post-abortion, post-partum, as well as those utilizing laparoscopic and mini-laparotomy techniques. We excluded the number of complications due to insufficient details on the types of complications and their associated costs. The HMIS data was assessed through the National Data and Analytics Platform (NDAP). (8)

HMIS provided data on 736 districts, from which we excluded four that were not present in the 2019 reference shapefile: Malerkotla, Noklak, Mayiladuthurai, and Gaurella Pendra Marwahi. (9) Out of the remaining 732, we excluded 25 districts with no recorded tubectomies or vasectomies. A total of 707 districts were included in the study **(Figure 1)**. For these districts, we obtained the 2019 population of women and men in the reproductive age group from WorldPop. (10) The population was aggregated at the district, state, and national levels.

**Figure 1:**
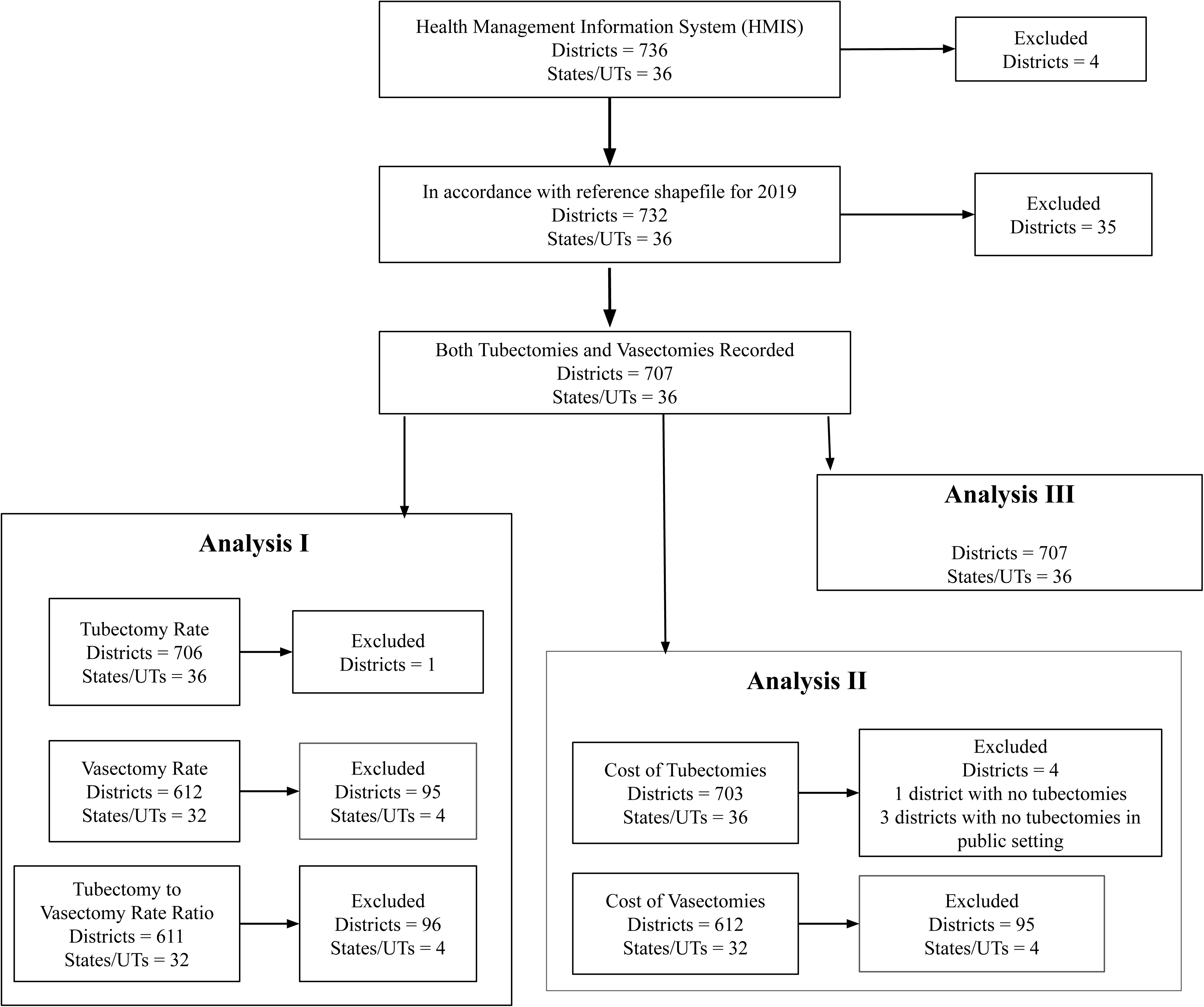
Analytic samples of districts and states/UTs.

The procedure costs of tubectomies were obtained from the Pradhan Mantri Jan Arogya Yojana (PMJAY) Health Benefits Package (HBP) 2022 **(Table 2)**. (11) The costs considered for analysis were different for laparoscopic and open tubal ligations, which include minilap, post-partum, and post-abortion tubal ligations. The costs of vasectomy procedures were obtained from a research study by Seamans and colleagues, as there were no estimates in recent official documents. (12) We used the cost of Ligation and Excision (L and E) vasectomy method from their work, as it is the most commonly performed method in India. (12)

**Table 2:** Costs of procedures and compensation for failures and deaths are considered in the analysis. All costs are adjusted to USD for the year 2022; 1 USD = 78.6 INR. All values are rounded to one decimal place for consistent presentation.

| **Procedure** | **INR**<br>**(2022)** | **USD**<br>**(2022)** |
| --- | --- | --- |
| Open tubal ligation (includes post-abortion, post-partum, and mini-laparoscopic) | 7000 | 89.1 |
| Laparoscopic tubal ligation | 10000 | 127.2 |
| Vasectomy (Ligation and excision) | 1987 | 25.3 |
| Compensation for sterilization failure | 30000 | 517.8 |
| Compensation for death following sterilization within 7 days | 200000 | 3452.3 |
| Compensation for death following sterilization within 7-30 days | 50000 | 863.1 |
| **For high-focus states** |  |  |
| Compensation for loss of daily wages, tubectomy | 2000 | 38.4 |
| Compensation for loss of daily wages, tubectomy, and postpartum sterilization | 3000 | 57.5 |
| Compensation for loss of daily wages, vasectomy | 2700 | 51.8 |
| **For non-high-focus states** |  |  |
| Compensation for loss of daily wages, tubectomy |  |  |
| - Below poverty line (BPL) or Scheduled Castes/Tribes (SC/ST) populations | 1000 | 19.2 |
| - Non-BPL or Non-SC/ST | 650 | 12.5 |
| Compensation for loss of daily wages, vasectomy | 1500 | 28.8 |

We used the Family Planning Indemnity Scheme to obtain the cost of compensation for failure and death resulting from the procedure. (13) For high-focus states/UTs, the government announced the enhanced compensation scheme in 2014. The cost of compensation for the loss of daily wages was taken from this scheme. (14) The National Family Health Survey was used to obtain the total fertility rates, which were used to classify the states/UTs into high-focus (total fertility rate ≥ 2.1) and non-high-focus states/UTs. (6)

### 2.2 Data Analysis

#### 2.2.1 Analysis I: Gender disparity in surgical sterilization in India

We assessed the disparity in surgical sterilization uptake by calculating the tubectomy-to-vasectomy ratio. First, we calculated the rates of tubectomies per 10,000 women in the reproductive age (RA), i.e., 15-49 years **(Equations 1a)** Similarly, we calculated the vasectomy rate per 10,000 men in the corresponding age group **(Equation 1b)**. Next, we calculated the ratio by dividing the tubectomy rate by the vasectomy rate. A ratio of 1 represents no gender disparity, and a ratio of >1 would indicate that the burden of sterilization falls disproportionately on women. One hundred and twenty districts and four states/UTs, namely Sikkim, Andaman and Nicobar, Lakshadweep, and Mizoram, had no vasectomies, so they were excluded from vasectomy rate calculations. We calculated vasectomy rates for 612 districts and 32 states/UTs. We included 706 districts and 36 states/UTs in calculating the tubectomy rate, as 26 districts with no tubectomies were excluded. There was only one district where vasectomies were done, but no tubectomies were performed. For ratio calculations, we excluded 121 districts and 4 states/UTs where neither tubectomies nor vasectomies were recorded. Hence, the rate ratio calculation includes 611 districts and 32 states/UTs **(Figure 1)**.

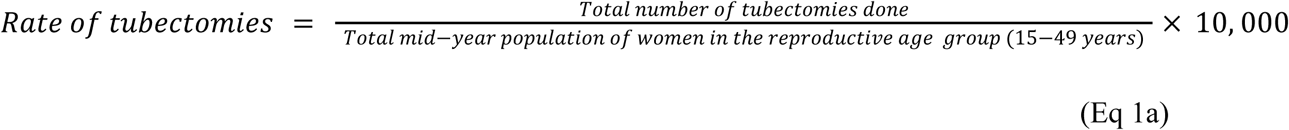

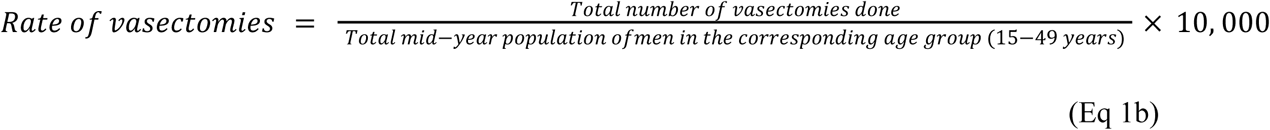

The failure rates for tubectomies and vasectomies were calculated as the number of failures per 10,000 procedures performed. Here, the number of failures post-tubectomies and vasectomies is defined as the occurrence of pregnancy after the procedure. (15) The reporting biases for failures are discussed ahead under study limitations. The failures and deaths were used in further analyses (see **Section 2.2.3** ahead). For post-tubectomy failures and deaths, we included 706 districts across 36 states/UTs, as 26 of 732 districts had no recorded tubectomies. For post-vasectomy failures and deaths, we included 612 districts and 32 states/UTs, as 120 districts and 4 states/UTs recorded no vasectomy **(Figure 1)**.

#### 2.2.2 Analysis II: Cost of surgical sterilization

To calculate costs, we considered only procedures performed in public health centers, as the costs of procedures in private centers are variable and unavailable in the public domain. Also, surgical sterilizations in India are predominantly conducted in public health centers. (16) We adjusted the vasectomy costs for inflation from 2007 to 2022. (12) The total costs of male and female sterilization procedures were calculated by multiplying the number of procedures of each type by the per-procedure cost. The calculations for the number of different tubectomy procedures (open and laparoscopic) are presented in the **Supplementary Methods.** Only one surgical method (ligation and excision) is considered for calculating **t**he per-procedure cost for vasectomy. **Table 2** enlists all per-procedure costs.

The Government of India classifies states/UTs as high-focus or non-high-focus based on a total fertility rate cut-off of 2.1 to ensure better implementation of health and developmental schemes in high-fertility states/UTs. (6,17) NFHS-5 noted five high-focus states/UTs with a total fertility rate ≥ 2.1, including Bihar, Jharkhand, Meghalaya, Manipur, and Uttar Pradesh. (18)

We included three kinds of compensation costs. First, we calculated the population-level cost of lost daily wages by multiplying the procedure counts by the per-procedure lost-wage compensation cost. The per-procedure compensation cost for lost wages was higher in the high-focus states/UTs for both tubal ligation and vasectomy. Additionally, in high-focus states/UTs, per-procedure compensation costs for lost daily wages from tubectomies were further bifurcated by the timing of the procedure. Greater compensation was provided when the procedure was performed postpartum. For non-high-focus states/UTs, the per-procedure compensation costs for lost daily wages from tubectomies were bifurcated by beneficiary income level and caste. Greater compensation is provided to people below the Poverty Line (BPL) who belong to the Scheduled Castes (SC) or the Scheduled Tribes (ST). Second, we calculated the cost of compensation in the event of procedure failure by multiplying the number of failures for each procedure by the per-failure cost. Third, we calculated the cost of death compensation by multiplying the number of deaths for each procedure by the per-death compensation cost. As per the Family Planning Indemnity Scheme, the cost of compensation due to death following the procedure varies depending on the time of occurrence of death. Greater compensation is provided if death occurs during the procedure or within seven days following the discharge from the hospital, compared to death within 7-30 days after sterilization. The per-procedure compensation costs are mentioned in **Table 2**.

We calculated the total costs of vasectomies and tubectomies using **Equation 2**. Finally, the total cost of sterilization by the public health system was computed as the sum of the costs of vasectomy and tubal ligation.

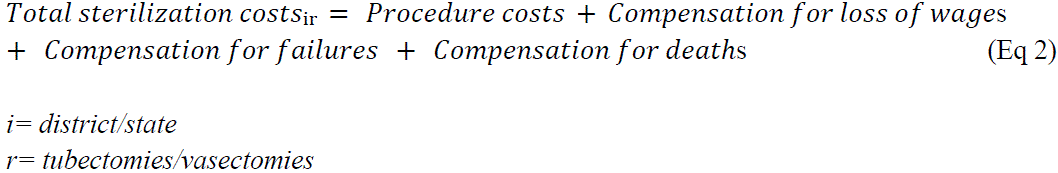

All costs were converted from INR to USD using the average 2022 exchange rate (1 USD = 78.593 INR). (19)

As HMIS did not provide deaths within and beyond seven days since the sterilization procedure or disaggregated data on sterilizations by caste and income groups, we calculated the minimum and maximum bounds to the total sterilization costs. There were two minimum and maximum cost calculations for tubectomies based on the type of state, i.e, high-focus or non-high-focus. For non-high-focus states/UTs, the minimum cost of tubectomies was calculated assuming all procedures were performed on women belonging to the non-BPL and non-SC/ST group. To the procedure cost and the cost of compensation for lost wages, we added the cost of compensation due to failure and the cost of compensation in the event of death within 7-30 days following sterilization. To calculate the maximum cost of tubectomies, we assumed that all procedures were performed on women in the BPL and SC/ST categories. The cost of failure following sterilization was similar to that described for the minimum-cost scenario. However, we added the cost of compensation, assuming death occurred within seven days of sterilization. For high-focus states/UTs, the maximum cost of tubectomies was calculated after adding costs of the procedure and compensation for postpartum and postabortion tubectomies.

To calculate the minimum cost of vasectomies, the procedure cost, compensation for lost wages, the cost of failure, and the cost of compensation were added, assuming death occurred within 7-30 days following sterilization. To estimate the maximum costs of vasectomies, the procedure cost, compensation for lost wages, cost of failure, and compensation costs were added, assuming all deaths occurred within seven days of sterilization.

The above calculations led to four scenarios for total costs: minimum cost of tubectomies and minimum cost of vasectomies; minimum cost of tubectomies and maximum cost of vasectomies; maximum cost of tubectomies and minimum cost of vasectomies; and maximum cost of tubectomies and maximum cost of vasectomies.

All district-, state-, and national-level costs are reported as ranges to show the minimum and maximum bounds. Of the 732 districts, we did not calculate the vasectomy costs for 120 because no vasectomies were recorded. We did not calculate tubectomy costs for 26 districts because no tubectomies were recorded, and 3 districts had no tubectomies performed in the public health system. Therefore, we calculated the costs of tubectomies for 703 districts and the cost of vasectomies for 612 districts. Among states/UTs, we included 36 in the tubectomy cost analysis. For vasectomy costs, 32 states/UTs were considered, as four states/UTs did not record any vasectomies. We included 706 districts for calculating compensation costs due to post-tubectomy failure or death, as 26 districts out of 732 had no tubectomies. For compensation costs due to post-vasectomy failure or death, we included 612 districts, as 120 districts recorded no vasectomy **(Figure 1)**. To illustrate the cost disparity, we calculated the percentage share of total vasectomy costs across 612 districts.

#### 2.2.3 Analysis III: Cost of surgical sterilization if gender equity is achieved

We estimated the total cost of surgical sterilizations if there was gender equity in India, i.e., if the rate of tubectomies was equal to the rate of vasectomies. As Analysis *III* relies on Analysis *II,* we considered procedures performed only in public health centers. Additionally, the percentages of different types of tubectomies, relative to the total number of tubectomies, remain unchanged. Ideally, in the absence of social pressures and skewed gender roles and responsibilities for family planning, there would be an equal acceptance of surgical sterilization for both men and women, resulting in comparable (near equal) rates across both. Therefore, our gender equity analysis assumed a benchmark vasectomy rate to be half of the total surgical sterilization rate. The benchmark rate was calculated as the average of the tubectomy and vasectomy rates. We calculated the new number of procedures based on the benchmark rates using **Equations 3a&b**.

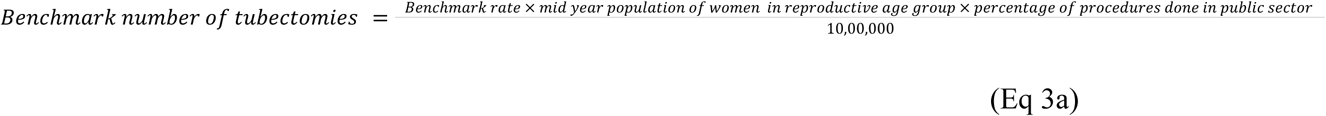

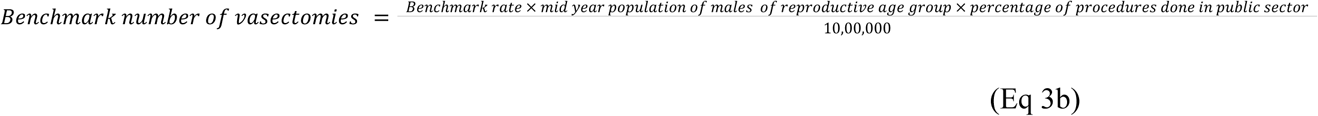

If no vasectomies were performed in a district, we assumed that at the new benchmark rate, they would be performed entirely within the public health system. There were 95 districts out of 707 in which we assumed that all the vasectomies would be performed in the public health system. There were 3 districts in which no tubectomies were performed in the public health system; we have assumed that all tubectomies will be performed as minilap sterilizations there, at benchmark rates. The benchmark number and benchmark costs were calculated for 707 districts for both tubectomies and vasectomies. The new number of failures following tubectomies and vasectomies was calculated by multiplying the failure rate (per 10000 population) calculated in *Analysis I* by the new number of procedures. The new death counts following tubectomy and vasectomy were also similarly calculated. The number of districts and states/UTs included for benchmark cost of compensation for failure and death was similar to that in *Analysis II* **(Figure 1)**. We estimated the new total costs of vasectomies and tubectomies under the gender-equity assumption, following the steps described in *Analysis II*.

The public health system’s savings were computed as the difference between the new total costs and the original total costs. At the district level, some districts showed positive savings, while the rest showed negative savings. For the state and national levels, we used two methods to estimate savings. First, we calculated net savings by summing the savings across districts, including both positive and negative values. Second, we calculated state– and national-level gross savings by summing only the districts with positive savings and excluding those with negative savings (i.e., losses). We report net and gross savings for India, 36 states/UTs, and 707 districts.

## 3 Results

### 3.1 Gender disparity in the uptake of surgical sterilization in India

Nationally, tubectomy and vasectomy rates were 96.5 per 10,000 women of RA and 1.4 per 10,000 men of RA, respectively, yielding a ratio of 67.5:1. Across states/UTs, tubectomy rates ranged from 5.6 per 10,000 in Sikkim to 185.8 per 10,000 in Puducherry. Chhattisgarh had the highest vasectomy rate of 8.3, while four states/UTs, including Andaman and Nicobar Islands, Lakshadweep, Mizoram, and Sikkim, recorded no vasectomies in 2019-20; All the remaining thirty-two states/UTs had a tubectomy-to-vasectomy rate ratio > 1, depicting a greater burden of surgical sterilization on women. Puducherry had the highest rate ratio of 758.4.

Among districts, Kheda in Gujarat had the highest tubectomy rate (357.6 per 10000 women of RA), while Gadchiroli in Maharashtra had the highest vasectomy rate (65.2 per 10000 men of RA). One hundred and twenty districts recorded no vasectomies. Of the remaining 612 districts, 611 have rate ratios >1, with Chittoor and Andhra Pradesh reporting the highest rate ratio of 6238.9:1 **(Figure 2)**.

**Figure 2:**
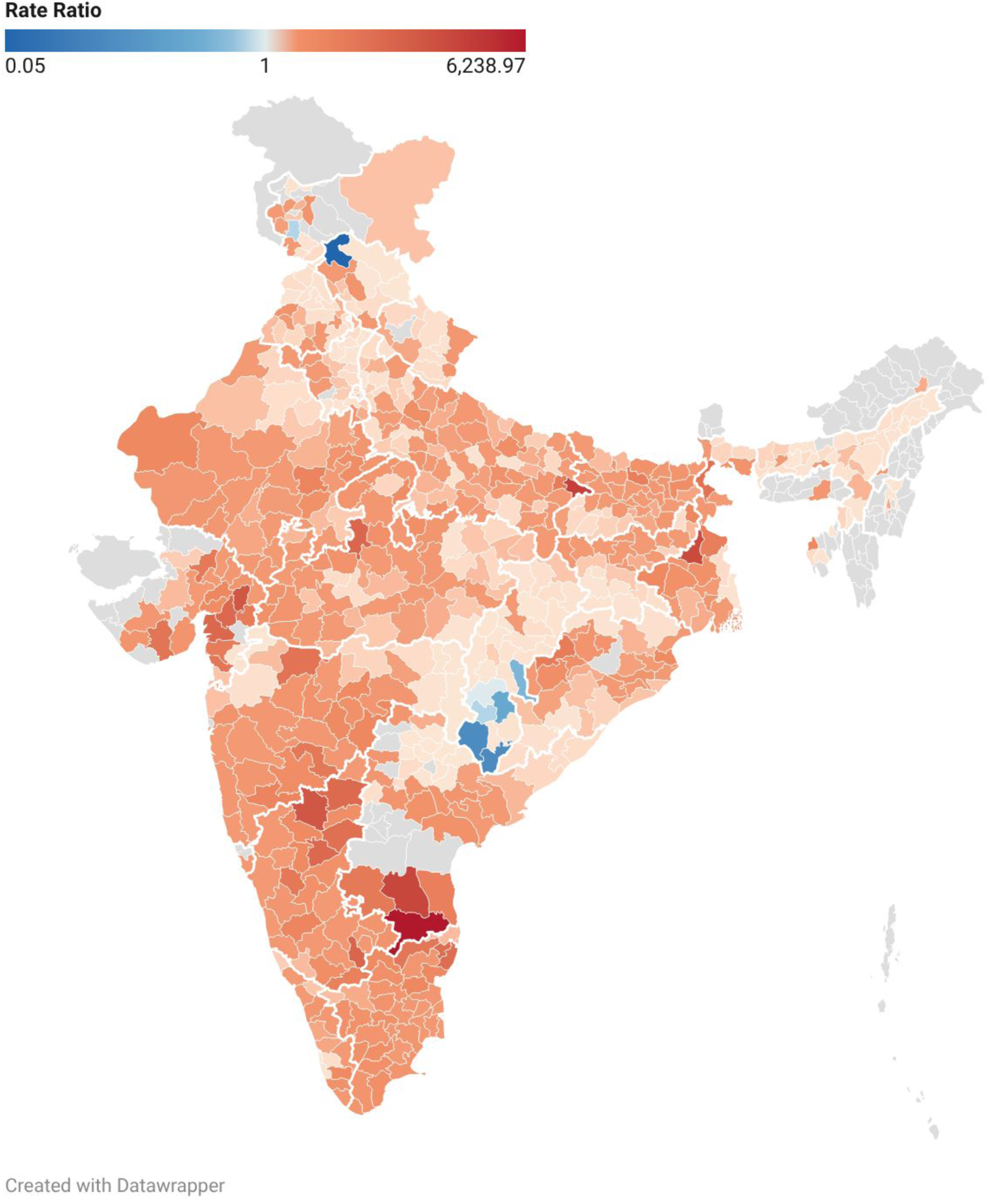
District-wise tubectomy-to-vasectomy rate ratio. The tubectomy-to-vasectomy rate ratios for 611 districts in India. Values centered at 1, which represents an equitable rate ratio. More red values indicate greater gender disparity against women. Draker shades of blue indicate greater vasectomy uptake than tubectomy uptake. The grey color denotes districts with missing data.

Nationally, the failure rate for tubectomy (6.6 per 10,000 tubectomies) was lower than that of vasectomy (13.5 per 10,000 vasectomies). Across states/UTs, Puducherry had the highest post-tubectomy failure rate of 48.7 per 10,000 tubectomies, while Meghalaya had the highest post-vasectomy failure rate of 2500 per 10,000 vasectomies, where only four procedures were performed, of which one failed. The highest failure rates among the districts were observed in Chamba, Himachal Pradesh, for tubectomies (526.3), and in Balangir, Odisha, for vasectomies (3333.3). However, it is noteworthy that HMIS recorded only three vasectomies in Balangir, Odisha, of which one failed. So, the rate value is inflated due to the small denominator.

### 3.2 Cost of surgical sterilization in India

#### 3.2.1. Total costs

Nationally, we estimated the total cost of surgical sterilization in public health centers to range from 305,254,127 to 313,982,589 USD. Of these, only 0.6-0.8% were due to vasectomies. Low total cost of vasectomies was also observed at the state and district levels **(Figure 3)**. Across states/UTs, vasectomy costs as a proportion of total costs ranged from 0% in four states/UTs (Andaman and Nicobar Islands, Mizoram, Lakshadweep, and Sikkim) to 5.7% in Chhattisgarh. There were 120 districts with no public health system costs for vasectomies. Bishnupur, Manipur, had the highest proportion of its costs on vasectomy (100%) among all districts.

**Figure 3:**
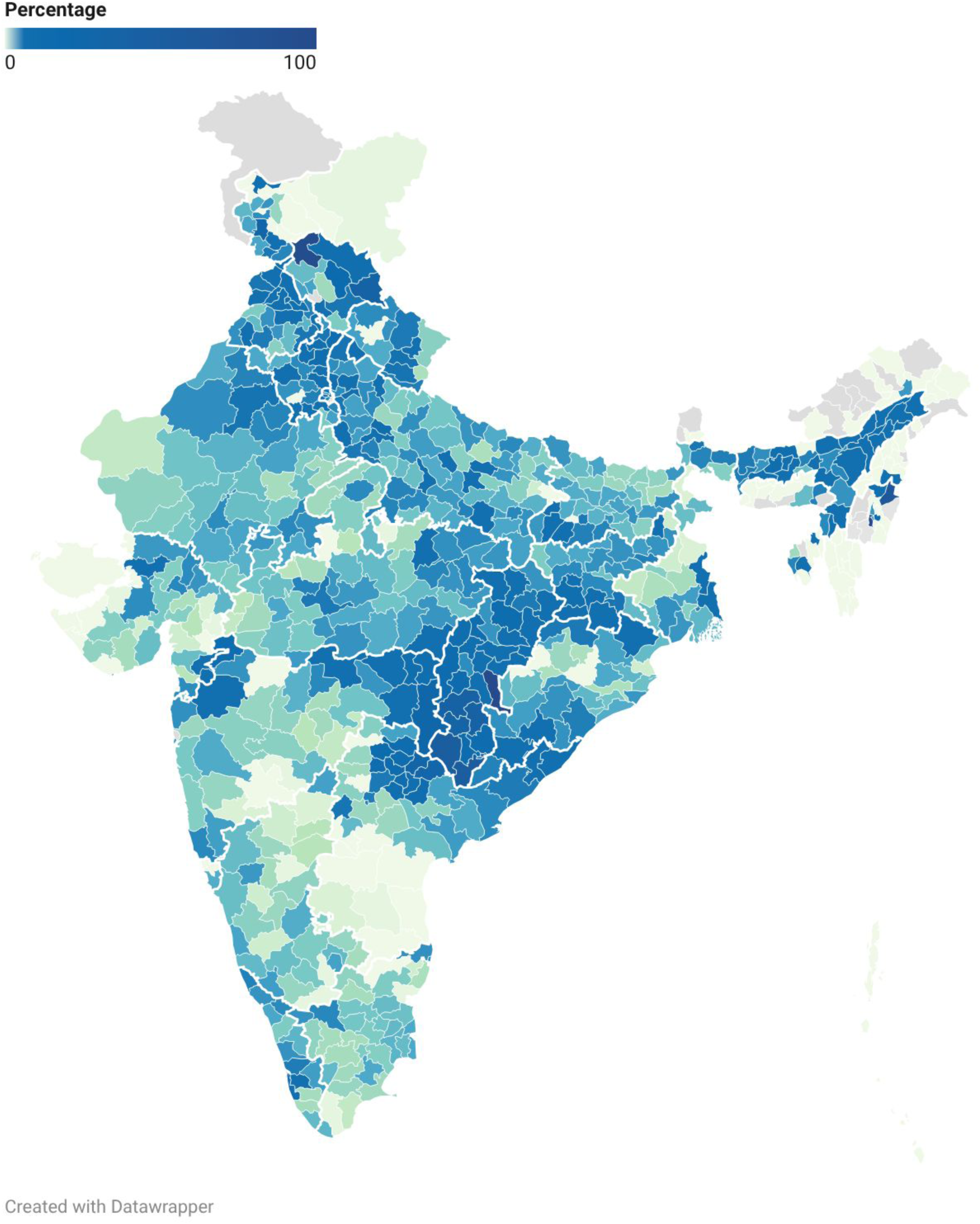
District-wise percentage of vasectomy costs relative to total surgical sterilization costs. The percentage of the total sterilization cost due to vasectomies. A total of 707 districts in India are colored by increasing intensity, indicating higher vasectomy spending. Grey color denotes areas with missing data.

Madhya Pradesh had the highest total tubectomy-related costs, ranging from 41,890,862 to 43,317,451 USD across different scenario analyses, while Maharashtra had the highest total vasectomy-related costs (387,737 USD). Sikkim and Lakshadweep (7,495 to 7,837 USD for each) had the lowest costs for tubectomies. Across districts, the highest costs for tubectomies were recorded in Ahmedabad, Gujarat (4,382,062 to 4,548,688 USD). Twenty-six districts have no tubectomies recorded and therefore have no associated tubectomy costs. Moreover, for 3 districts (Churachandpur, Kamjong, Pherzawl), we have no costs for tubectomies because no tubectomies were conducted in the public health system. Gadchiroli, Maharashtra, had the highest vasectomy costs (99,584 USD).

#### 3.2.2. Costs of procedures and compensation for lost wages

Nationally, the cost of the procedures and compensation for lost wages ranges from 304,660,129 to 313,300,777 USD. State-wise, the highest costs were noted in Madhya Pradesh (41,629,153 to 43,053,834 USD) for tubectomy and Maharashtra (379,339 USD) for Vasectomy. District-wise, the highest cost was noted in Ahmadabad, Gujarat (4,380,535 to 4,547,160 USD) for tubectomy and Gadchiroli, Maharashtra (97,563 USD) for vasectomy.

#### 3.2.3. Compensation costs for procedure failures

Nationally, the cost of compensation for tubectomy failures was 31 times that for vasectomy failures (863,817 USD vs. 27,865 USD). Maharashtra had the highest costs, that is, 208,033 USD, compensating for 545 tubectomy failures, and 8,398 USD for 22 vasectomy failures. Among districts, Buldhana, Maharashtra, had the highest compensation costs for tubectomy failures (22,139 USD), while Bhandara, Maharashtra, had the highest compensation costs for vasectomy failures (3,054 USD).

#### 3.2.4. Compensation costs for post-operative deaths

Nationally, the cost of compensation for deaths following tubectomies ranged from 28,628 to 114,514 USD, while that for vasectomies ranged from 636 to 2544 USD. Thus, the total compensation for deaths following tubectomies was about 45 times that of vasectomies. Maharashtra (8,270 to 33,082 USD) and Assam (636 to 2,544 USD) had the highest compensation costs for deaths following tubectomies and vasectomies, respectively. Among districts, Lawngtlai in Mizoram had the highest compensation costs for deaths from tubectomies (3,181 to 12,724 USD), and Nalbari in Assam had the highest compensation costs for deaths from vasectomies (636 to 2,545 USD).

There are minimum and maximum costs of tubectomies at the national level, considering two different death scenarios and different compensation rates for BPL and SC/ST **(Figure 4a)**. The minimum and maximum costs of vasectomies are shown in **Figure 4b**. All costs at the national and state levels are presented as ranges, reflecting the four scenarios. The different costs across scenarios are shown in **Figure 4c**.

**Figure 4:**
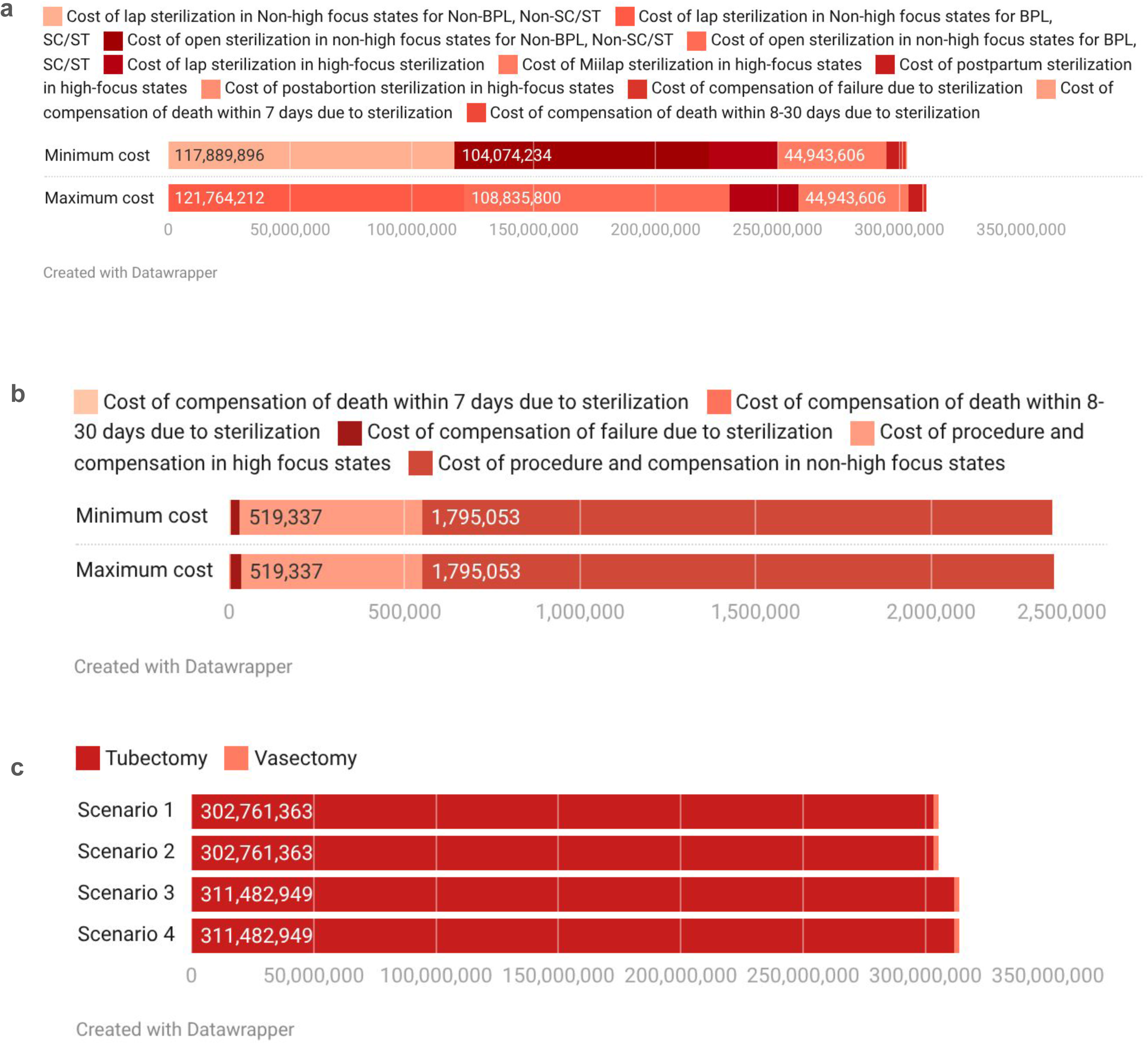
Minimum and maximum costs of a) tubectomies, b) vasectomies, and c) total sterilizations. Minimum cost of tubectomies = Cost of laparoscopic sterilization procedure and compensation assuming all belongs to Non-BPL, Non-SC/ST in Non-high focus states + cost of open sterilization assuming all belongs to Non-BPL, Non-SC/ST in Non-high focus states + cost of laparoscopic sterilization procedure and compensation if all values are assumed to be at the level of high focus states + cost of open sterilization and compensation if all values are assumed to be at the level of high-focus states + cost of postpartum sterilization and compensation if all values are assumed to be at the level of high focus states + cost of postabortion sterilization and compensation if all values are assumed to be at the level of high-focus states + cost of compensation assuming failure occurred following sterilization + cost of compensation assuming death occurred following sterilization within 8-30 days. Maximum cost of tubectomies = Cost of laparoscopic sterilization procedure and compensation assuming all belong BPL, SC/ST in non-high focus states + cost of open sterilization and compensation assuming all belong to BPL, SC/ST in non-high focus states + cost of laparoscopic sterilization procedure and compensation if all values are assumed to be at the level of high-focus states + cost of open sterilization and compensation if all values are assumed to be at the level of high-focus states + cost of postpartum sterilization and compensation if all values are assumed to be at the level of high-focus states + cost of postabortion sterilization and compensation if all values are assumed to be at the level of high-focus states + cost of compensation assuming that failure occured following sterilization + cost of compensation assuming death occurred following sterilization within 7 days. Minimum cost of vasectomies = Cost of procedure and compensation if all values are assumed to be at the level of non-high focus states + cost of procedure and compensation if all values are assumed to be at the level of high-focus states + cost of compensation assuming that failure occurred following sterilization + cost of compensation assuming death occurred following sterilization within 7-30 days. Maximum cost of vasectomies = Cost of procedure and compensation if all values are assumed to be at the level of non-high focus states + cost of procedure and compensation if all values are assumed to be at the level of high-focus states + cost of compensation assuming that failure occurred following sterilization + cost of compensation assuming death occurred following sterilization within 7 days. Cost of sterilization was calculated across four scenarios: Scenario 1 = Minimum cost of tubectomy + Minimum cost of vasectomy; Scenario 2 = Minimum cost of tubectomy + Maximum cost of vasectomy; Scenario 3 = Maximum cost of tubectomy + Minimum cost of vasectomy; Scenario 4 = Maximum cost of tubectomy + Maximum cost of vasectomy. National costs are written in these figures, which include 36 states and 707 districts.

### 3.3 Estimated cost of surgical sterilization in India if there were gender equity in uptake

The new number of vasectomies, based on the benchmark rate (48.9 per 10,000 men of RA), was 1,873,384, a gain of 1,818,669 vasectomies nationally. The state requiring the largest increase in vasectomy numbers to achieve the equity benchmark was Karnataka (181,540). The district requiring the largest increase was Surat, Gujarat (43,072). Similarly, the number of tubectomies, based on the benchmark rate, was calculated to be 1,722,288 about 1,672,623 fewer than the original estimate.

The total cost of surgical sterilization, if the equitable benchmark rate is achieved, was estimated at 238,465,812 to 242,904,608 USD, resulting in net savings of 62,349,519 to 75,516,777 USD. Nationally, gross savings would range from 69,343,449 to 81,450,411 USD. A total of 34 states/UTs were estimated to have net financial benefits ranging from 1,629 to 2,144 USD for Sikkim, to 12,239,744 to 14,388,098 USD for Madhya Pradesh **(Figure 5a)**. Manipur and Mizoram were estimated to have incurred losses of 455,838 and 32,690 USD, respectively. The state with the highest gross savings is Madhya Pradesh (12,270,080 to 14,404,667 USD), and the lowest is Sikkim (1,629 to 2,144 USD). Of the 707 districts, 635 noted positive net savings. Gross savings are noted in 635 out of 707 districts. The district with the highest net and gross savings would be Ahmadabad, Gujarat (1,393,904 to 1,644,214 USD). The gross savings of all the states/UTs are shown in **Figure 5b**. **Figure 6a** shows districts with positive and negative savings included in the net saving estimates at the state and national levels. **Figure 6b** shows only the districts with positive savings that contributed to the gross savings at the state and national levels.

**Figure 5:**
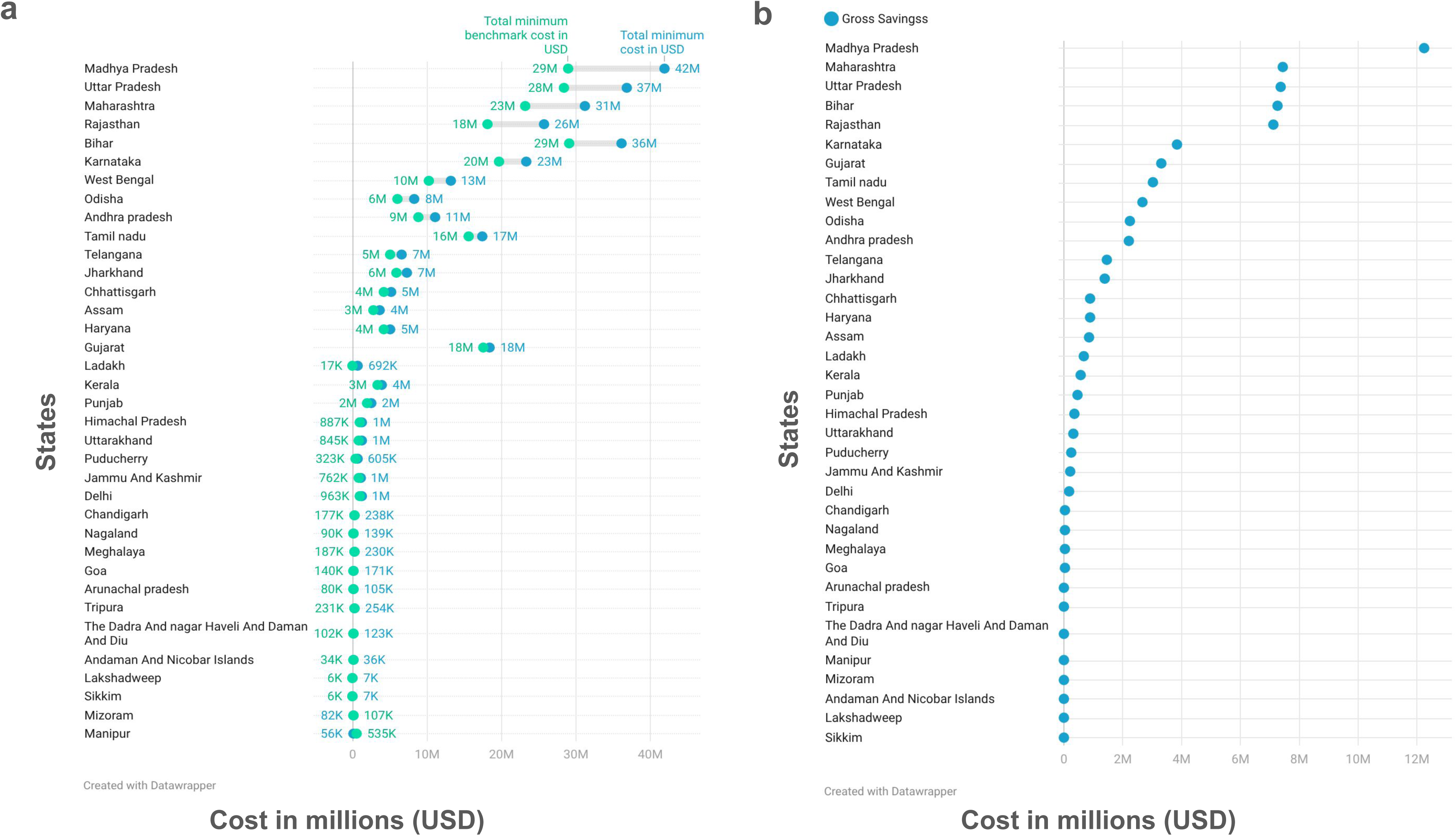
State-wise a) net and b) gross savings after vasectomy scale-up. The state-wise net savings after scaling up the vasectomy rate to match the tubectomy rate, assuming the total sterilization rate to be constant. Green dots indicate the minimum sterilization cost, whereas blue dots indicate the minimum cost after scaling up vasectomy rates. Numbers with the plus sign denote positive savings for the states, after scaling up vasectomies. Numbers with a minus sign denote net losses to the states. Gross savings increase as the blue dots move to the right. These are state-wise gross savings after scaling up the vasectomy rate to match the tubectomy rate, assuming the total sterilization rate to be constant. All the costs are in USD 2022. We included all 36 states in the figures.

**Figure 6:**
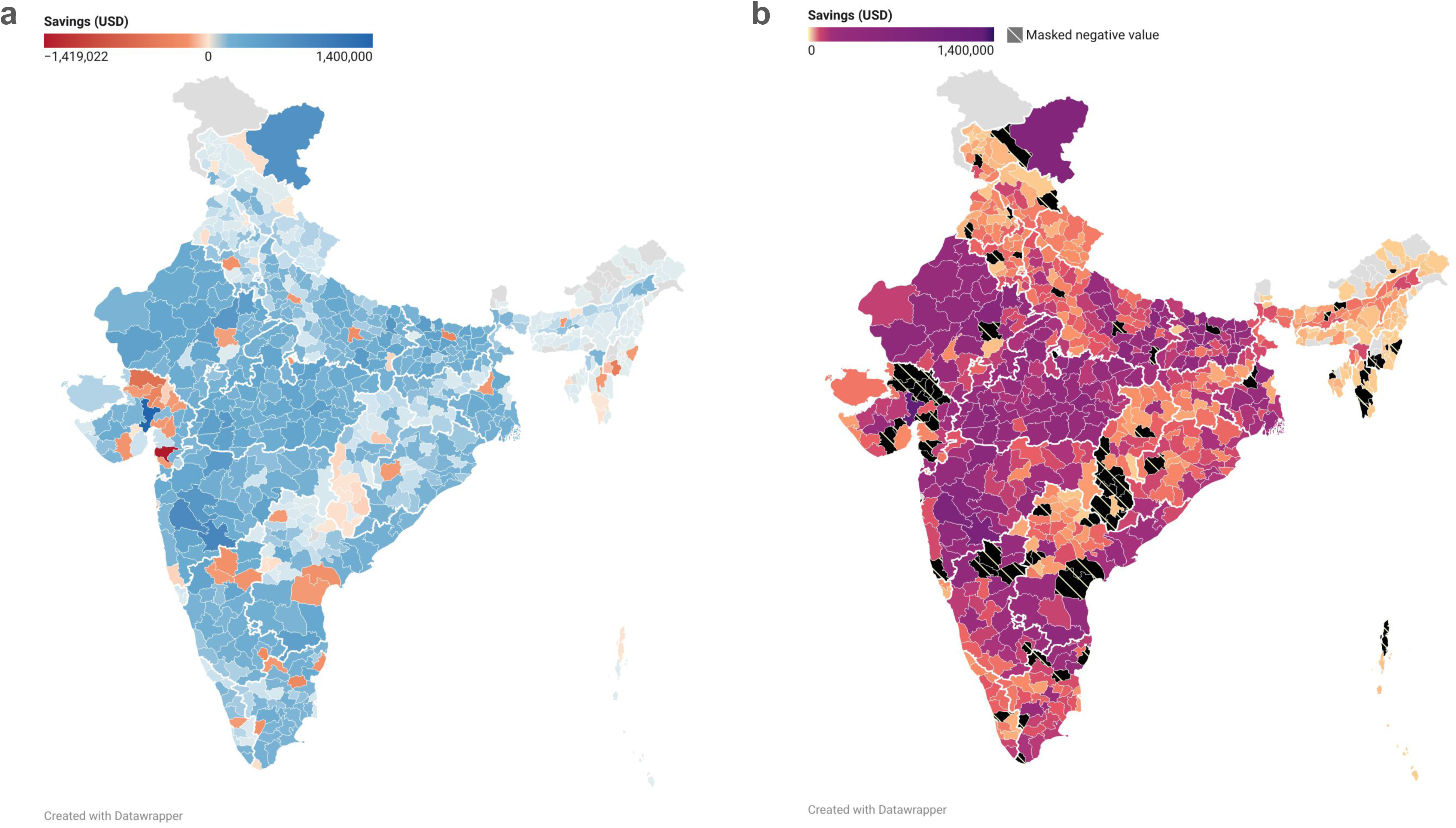
a) Districts with positive and negative savings included in the net saving estimates at the state and national levels, and b) The districts with positive savings that contributed to the gross savings at the state and national levels. Net savings increase as the color gradient changes from yellow to violet. The grey color denotes districts with missing data. District-wise gross savings from surgical sterilization if vasectomies are scaled up to match the tubectomy rate, while keeping the total sterilization rate constant. The districts with negative savings are masked in black. Gross savings increase as the color gradient changes from yellow to violet. All the costs are in USD 2022. We included 707 districts in the figures.

## 4 Discussion

### 4.1 Summary of Findings

Our study highlights the disparity in the uptake of surgical sterilization between Indian women and men. The rate of tubectomies was 67.5 times higher than that of vasectomies. Of the 32 states and union territories analyzed for vasectomies, 22 had a vasectomy rate under 1 per 10,000 men of reproductive age. The rate ratio of tubectomies to vasectomies was skewed toward women (>1) in 603 districts and 32 (88.8%) states/UTs.

Surgical sterlizations costed 305 to 314 million USD to the Indian public health system in 2019-20, of which 99.2-99.4% costs were for tubectomies. The cost of compensation for tubectomy failures was 31 times that of vasectomy failures (863,817 USD vs. 27,865 USD), and the cost of compensation for the death following tubectomies was about 45 times that of vasectomies across multiple analysis scenarios. We estimated that the public health system will save over 62-75 million USD if vasectomies are scaled up to achieve gender-equitable rates.

### 4.2 Reasons for low vasectomy utilization

There are several historical, sociopolitical, and cultural reasons for gender-based disparity in surgical sterilization rates and low uptake of vasectomies in India. While a comprehensive discussion is beyond the scope of the current article, below we discuss some reasons for low vasectomy uptake.

In 1952, India became the first country to implement a nationwide family planning program to control birth rates and stabilize the population. Following up, the first vasectomy program was launched in 1954. (20) During the years of political Emergency (1975 to 1977), the program took a coercive turn, mandating sterilization in certain parts of the country. The total number of vasectomies increased from 0.61 million in 1974-75 to 1.3 million in 1975 and 8.1 million in 1977. (21,22) A sharp decline was reported following the Emergency. (21) The extensive opposition to coercion during the Emergency contributed to the historical defeat of the ruling party in 1977. (23) However, the enforcement of the vasectomy program during the Emergency led to a distrust of family planning policies, especially among men, and a hesitation on the part of program implementers to target men in family planning efforts at the community level, which arguably contributed to the disparity over decades. (23,24)

Gender inequity in surgical sterilizations was first recognized in the National Population Policy (NPP) in 2000. Improving awareness among men about vasectomy as a safe and straightforward procedure was considered a solution to increase uptake among men. (25) Several cross-sectional studies across different parts of India have shown that the awareness about vasectomy in men and women is high (70-81.5%). (26–29) The low service utilization despite high awareness exposes the possible role played by patriarchal norms about sterilization among men, shaped by sociocultural, religious, and economic factors. (24)

Various studies have demonstrated misconceptions among men and women regarding vasectomies, influenced by patriarchal gender norms. These include misconceptions among men regarding vasectomies, such as loss of sexual pleasure, physical weakness, and loss of livelihood. (30–32) The misconceptions are not exclusive to men. Women, too, are known to believe that a vasectomy will affect the work performance of their husbands and affect their family income. (33) Additionally, in multi-generational households that are common in India, the choice of method and timing of contraception is influenced by mothers-in-law and other members of the household and extended family, putting more pressure on the women to bear the burden of contraception use. (34,35) Evidence beyond India also shows that gender disparity and surgical sterilization are closely linked. Work in Costa Rica reported that men who had more gender-equitable views were more likely to undergo vasectomies. (36) Similarly, it has been seen that countries that have high (measured) gender equality also rank high in vasectomy rates. As per the adjusted Gender Inequality Index, eight of the top ten countries also feature in the top ten countries with the highest vasectomy rates. (37) These highlight the role of existing gender norms and patriarchy in the problematic conceptualization of vasectomy among communities with policy implications.

The relatively low uptake of vasectomies compared to tubectomies can also be traced back to family planning being framed as a ‘women’s issue’. (38) Historically, a focus of programs such as the National Health Mission was to improve maternal and child health indicators. Consequently, India’s family welfare programs target women as the primary beneficiaries. Contraception is seen as a necessity to reduce maternal and child mortality. (24) Out of the eight family planning methods promoted by the Government in India, six focus on women. (39) The disproportionate focus on and uptake of contraceptive methods for women is highlighted by the National Family Health Survey. In 2019-21, among married women aged 15-49 years, 38% used tubectomies, 5% used Oral Contraceptive Pills (OCPs), 2.1% used intrauterine devices, and 10% used traditional methods such as the rhythm and withdrawal methods. Among the married men, 68.5% did not use any kind of contraceptive, 5.7% used condoms, and only 0.3% of men underwent vasectomy. (6) The yearly target for tubectomies is eight to ten times higher than that of vasectomies across health centers in the country. (24) This specific focus of these vertically structured programs on women renders contraception a necessity for women but a choice for men.

Additionally, frontline healthcare workers in India, including Accredited Social Health Activists (ASHAs), are primarily women. Their outreach activities coincide with men’s working hours, which provides fewer opportunities for direct engagement. (24) Also, men who want to be involved in family planning decisions and learn about sexual and reproductive health have reported preferring a male provider to discuss their issues. (40,41) Women healthcare workers have also reported facing verbal abuse from men during outreach efforts about sexual and reproductive health, in part due to patriarchal gender norms and restricted societal conventions. Consequently, women health workers prefer discussing vasectomy in the presence of healthcare workers who are men. (24) However, men in healthcare are few at the grassroots level and not actively engaged in outreach efforts. Therefore, the lack of community health workers who are men also creates a barrier for men who wish to learn more about sexual and reproductive health. (42)

### 4.3 Policy implications and recommendations

We estimate that the Indian public health system would save at least 62 million USD if vasectomy rates increased to a gender-equitable benchmark. Beyond economic benefits, given that vasectomies are simpler and safer than tubectomies, increased rates of vasectomies will reduce the total number of deaths following sterilization by reducing the post-tubectomy deaths.

India’s family planning policies and programs need to account for and actively work to remove the barriers rooted in structural factors, such as patriarchal gender norms, to the low vasectomy uptake. At present, the Information, Education, and Communication (IEC) materials available to frontline healthcare workers have been reported to only focus on providing information, instead of addressing gender norms and negative perceptions of masculinity associated with vasectomy. (24) Improving the content and quality of IEC, along with mass media campaigning, can assist frontline healthcare workers in increasing vasectomy uptake. For instance, a Brazilian mass media campaign promoting vasectomies was associated with a 108% increase in uptake. Further, periodic mass media campaigns alleviated the long-term downward trend in vasectomies over 12 years, indicating sustained improvements. (43)

Additionally, men in healthcare should be employed and trained to carry out outreach activities. Outreach activities that are conducive to men’s engagement, such as those in workplaces, can help increase vasectomy uptake. These activities should aim to challenge gender norms actively, increasing the participation of men in broader family planning and their outlook towards sexual and reproductive health. In a study conducted across nine districts of Uttar Pradesh, male healthcare workers communicated with men about no-scalpel vasectomy (NSV) and addressed their queries. Female healthcare workers communicated with their spouses. Over two years, the number of NSVs increased threefold in these districts. (31)

There is a need to shift from a target-based population-control agenda to a target-free client-based approach. Target-based approaches are inherently coercive. While India’s family planning policy is advertised as a client-based approach, states are allowed to set targets to achieve family planning goals. (24) Hence, female sterilization may be the most popular contraceptive method. Yet, it might not be due to any popular demand. It has been reported that women are often not offered or educated about alternative contraception options, making tubectomies the default. (44) Learning from the experience during the Emergency years, the focus must be shifted towards an educational and rights-based approach. For instance, in a study in Davao City, the Philippines, culturally sensitive information campaigns and couple-focused counseling led to an annual 80% increase in the number of NSV clients. (45) Similarly, a program facilitating client-centric communication on NSV led to a threefold increase in the total number of vasectomies. (46) Client-centric counseling can lead to increased participation of men in family planning, increasing vasectomy uptake.

Thus, regardless of the specifics of the intervention, our rationale is that the savings could be used for programs that improve gender equity in surgical sterilization uptake.

### 4.4 Limitations and Strengths

This study has several limitations. First, we excluded procedures performed in the private health sector from cost calculations because reliable, consistent cost data for these centers were unavailable. However, nearly three-fourths of the surgical sterilizations were performed in public health centers in India. (16) Moreover, the cost of the procedures in private centers is unregulated and can vary across centers. Furthermore, compensation for the loss of daily wages is provided only at public health centers. Therefore, excluding procedures performed in the private sector will have a negligible impact. Second, we excluded costs associated with post-procedure complications, as these can vary by type and severity. Such granular data on complications is unavailable in HMIS. Third, more generally, our analysis inherits the limitations of the HMIS as a data source, including facility coverage, data completeness, and reporting accuracy. (47) This issue can be more relevant to small districts where disentangling the noise from real data can be challenging. Fourth, there have been concerns about HMIS underreporting the number of failures of tubectomies and vasectomies. As per the Family Planning Division of the Ministry of Health and Family Welfare, the failure rate of minilap tubectomy and laparoscopic tubal occlusion was 5 per 1,000 procedures, whereas the national failure rate for tubectomies we calculated was 7 per 10,000. (48) Even so, extracting the HMIS data from the NDAP ensured that we only consider the data that passes the basic sanity checks for completeness described elsewhere in the NDAP documentation. (7) Fifth, our analysis used 2019-20 data, as the most recent HMIS data for district-level indicators are not publicly available. However, using 2019-20 data provides an important pre-COVID baseline for comparison with future studies. Sixth, the number of sterilization failures is a recall variable, which limits its credibility. Future studies can conduct more comprehensive bias analyses to assess the impact of bias. Lastly, due to the lack of publicly available data on vasectomy costs, the costs were taken from a paper that modeled the cost-effectiveness of different vasectomy methods. (12)

Despite these limitations, the study also has strengths. This is the first-of-its-kind pan-India analysis that highlights gender disparity in surgical sterilization uptake. We reported disparities in surgical sterilization using comparable rate measures across places and times. We also investigated the associated costs of surgical sterilization at the national, state, and district levels, thereby increasing the granularity of the findings for local decision-making. The choice of HMIS, a government data source, was deliberate to help policymakers make data-driven decisions using information that is easily and routinely available to them. Another strength of our study is the conceptualization and application of a benchmark that provides an empirical reference point for gender equity.

## 5 Conclusion

Increasing the number of vasectomies to achieve gender equity in surgical sterilization rates will save India money and reduce post-procedure mortality. India’s family planning policies and programs need to be responsive to the effects of patriarchal gender norms responsible for low vasectomy uptake. Improving the quality of educational and awareness materials and training more frontline male healthcare workers can help increase vasectomy uptake. A rights-based educational approach that emphasizes couple-centric counseling will ensure greater engagement of men in family planning. Future research should examine the effects of such interventions on increases in vasectomy uptake. Such research could use the disparity and cost measures used in this study for monitoring and evaluation.

## Declarations

### Ethics approval and consent to participate

Not applicable. We used publicly available aggregated data for this research.

### Consent for publication

Not applicable

### AI use

We did not use any generative AI tool to draft this manuscript. We used Grammarly for proofreading.

## Authors’ contributions

### Study concept and design

Siddhesh Zadey, Sharvari Mande, Anoushka Arora, Parth Sharma

### Acquisition, analysis, or interpretation of data

Sharvari Mande, Anoushka Arora, Siddhesh Zadey, Parth Sharma

### Drafting of the manuscript

Sharvari Mande, Anoushka Arora, Varun Raj Passi, Aiman Perween Afsar, Siddhesh Zadey, Parth Sharma

### Critical revision of the manuscript for important intellectual content

All authors

### Statistical analysis

Sharvari Mande, Anoushka Arora

### Administrative, technical, or material support

Siddhesh Zadey, Parth Sharma

### Study supervision

Siddhesh Zadey

## Dataset Availability

Data used and generated in this manuscript are available in the associated repository on Harvard Dataverse (https://doi.org/10.7910/DVN/OYUJIZ).

## Data Availability

Data used and generated in this manuscript are available in the associated repository on Harvard Dataverse https://doi.org/10.7910/DVN/OYUJIZ

