## Supplement for "Estimating gender disparities in surgical sterilization uptake in India in 2019-20 and cost savings from equity achievement"

### *1.1 Calculating the number of procedures conducted in public institutions*

To calculate the costs of surgical sterilizations in Analysis II, we considered procedures performed only in public institutions. The total costs of male and female sterilization procedures were calculated by multiplying the number of procedures of each type by the per-procedure cost. Since we did not have data on the number of tubectomies taking place post-abortion, post-partum, as well as those utilizing laparoscopic and mini-laparotomy techniques in public institutions, we calculated it using data from the Health Management Information System 2019-20.

First, we calculated the number of laparoscopic procedures taking place in both private and public institutions using **Equation 1**.

$$\text{Laparoscopic procedures in public \& private institutions} = \frac{(\text{Laparoscopic sterilizations to total sterilizations \%})\times(\text{Total sterilizations conducted in private and public institutions})}{100}$$

(Eq 1)

Next, we calculated the number of laparoscopic procedures conducted in public institutions using **Equation 2.**

$$\text{Laparoscopic procedures in public institutions} = \frac{(\text{Laparoscopic sterilizations at public institutions to total laparoscopic sterilisations \%})\times(\text{Laparascopic sterilizations in public \& private institutions})}{100}$$

(Eq 2)

Equations 1 and 2 were similarly used to calculate the number of tubectomies taking place post-abortion, post-partum, as well as those utilizing mini-laparotomy techniques.

### *1.2 Calculating the range of total sterilization costs*

We calculated a range of total sterilization costs at the district/state level for tubectomies/vasectomies. We hypothesized two scenarios that correspond to the minimum and maximum costs, thereby defining the lower and upper bounds of the range. For high-focus states, the minimum cost spent by the government would be when all deaths occurred within 7-30 days. The maximum cost would be incurred when all deaths occurred within 7 days. Refer to **Equations 3 and 4**.

$$\text{Minimum total sterilization costs}_{\text{ir}} = \text{Procedure costs} + \text{Compensation for loss of daily wages} + \text{Compensation for failures} + \text{Compensation for deaths occurring within } 7-30 \text{ days}$$

(Eq 3)

$$\text{Maximum total sterilization costs}_{\text{ir}} = \text{Procedure costs} + \text{Compensation for loss of daily wages} + \text{Compensation for failures} + \text{Compensation for deaths occurring within } 7 \text{ days}$$

(Eq 4)

*i= district/state*
*r= tubectomies/vasectomies*

Similarly, we hypothesized two scenarios for non-high-focus states. The minimum cost spent by the government would be when all deaths occurred within 7-30 days, and all patients were not below the Poverty Line (BPL) and/or did not belong to a Scheduled Caste (SC) or a Scheduled Tribe (ST). The maximum cost would be incurred when all deaths occurred within 7 days, and all patients were BPL and/or SC/ST. Refer to **Equations 5 and 6**.

$$Minimum\ total\ sterilization\ costs_{\mathrm{ir}} = Procedure\ costs + Compensation\ for\ loss\ of\ daily\ wages + Compensation\ for\ failures + Compensation\ for\ deaths\ occurring\ within\ 7 - 30\ days \quad \text{(Eq 5)}$$

$$Maximum\ total\ sterilization\ costs_{\mathrm{ir}} = Procedure\ costs + Compensation\ for\ loss\ of\ daily\ wages + Compensation\ for\ failures + Compensation\ for\ deaths\ occurring\ within\ 7\ days \quad \text{(Eq 6)}$$

*i= district/state*
*r= tubectomies/vasectomie*
*m=not BPL and/or SC/ST*
*n=BPL and/or SC/ST*

We are aware that other scenarios can occur, for example, the patient was BPL and death occurred within 7-30 days. However, the total cost of the procedure would fall within the upper and the lower bounds. Hence, costs excluding the maximum and minimum costs were not calculated.
